# Systematic review of health literacy champions: Who, what and how?

**DOI:** 10.1101/2023.03.15.23287331

**Authors:** Julie Ayre, Michael Zhang, Dana Mouwad, Dipti Zachariah, Kirsten McCaffery, Danielle Muscat

## Abstract

**Background:** Health literacy is an important aspect of equitable, safe, and high-quality care. For organisations implementing health literacy initiatives, using ‘change champions’ appears to be a promising strategy. This systematic review aimed to identify the empirical and conceptual research that exists about health literacy champions.

**Methods:** We conducted a systematic literature search using MEDLINE, Embase, CINAHL, Scopus, and PubMed, with additional studies identified by searching references and citations of included studies and reviews of organisational health literacy.

**Results:** Seventeen articles were included in the final review (case studies, n=9; qualitative research, n=4; quasi-experimental, n=2; opinion articles without case studies, n=2). Most articles had a high risk of bias. Often champions were not the focus of the article. Champions included staff across frontline, management and executive levels. Only five studies described training for champions. Key champion activities related to either 1) increasing organisational awareness and commitment to health literacy, or 2) influencing organisational strategic and operational planning. The most common output was ensuring that the organisation’s health information materials met health literacy guidelines.

Articles recommended engaging multiple champions at varying levels within the organisation, including the executive level. Limited funding and resources were key barriers. Three of five articles reported positive impacts of champions on implementation of health literacy initiatives.

**Discussions:** Few articles described health literacy champions in adequate detail. More comprehensive reporting on this implementation strategy and further experimental and process evaluation research are needed to progress this area of research.

**Registration:** This systematic review was registered with PROSPERO (CRD42022348816)

**Lay summary:** *Why did we do this review?:* Health organisations may want to improve their health literacy practices. Studies suggest that ‘champions’ may help bring about change in an organisation.

*What did we do?:* We searched five research databases to find out what research exists about health literacy champions.

*What did we find?:* We found seventeen relevant articles. Most had a high risk of bias. Often champions were not the focus. Champions could be staff across all levels (frontline, management, executive). Only five studies described training. Champions had two main types of activities: 1) making staff more aware and committed to health literacy; 2) changing organisational strategies and processes. Often this meant making sure that health information met health literacy guidelines. The articles suggested it may be better to have more than one champion, at different levels within the organisation, including the executive level. Three of five articles reported that champions may have improved health literacy practices.

*What does this mean?:* Research does not tell us much about health literacy champions or their impact on health literacy practices. We need studies that describe champions and their training in detail, and test their impact on health literacy practices at different sites.

---

Health literacy is an important consideration for any health organisation that seeks to provide equitable, safe, and high-quality care. This is clearly demonstrated across a range of health outcomes: low health literacy is associated with higher mortality, morbidity, medication errors, and rates of hospitalisation and emergency department visits (Berkman et al., 2011). Though these associations relate to an individual’s health literacy (i.e. skills to access, understand, appraise, and use health information and services), we must recognise the critical role that health organisations also play (Nutbeam & Muscat, 2021). For example, organisational structures and resources affect how easily people can navigate a health service, the quality of health information provided to patients, and extent that staff are trained in health literacy concepts and communication skills (Farmanova et al., 2018).

For organisations implementing health literacy initiatives, using ‘change champions’ appears to be a promising strategy. The Consolidated Framework for Implementation Research (CFIR) defines champions as “*individuals who dedicate themselves to supporting, marketing, and ‘driving through an [implementation]’, overcoming indifference or resistance that the intervention may provoke in an organization*”(CFIR Research Team-Center for Clinical Management Research, 2022; Greenhalgh et al., 2004). A recent scoping review identified change champions as one of four critical factors for implementing organisational health literacy interventions (Kaper et al., 2021). Similarly, a 2018 systematic review on the same topic identified the absence of a change champion as one of 13 key barriers (Farmanova et al., 2018). These findings reflect broader healthcare implementation research. For example, reviews show ‘generally positive’ evidence that champions contribute meaningfully to implementation efforts, and implementation science experts consider ‘identifying and preparing champions’ an important and highly feasible implementation strategy that should be prioritised (Lennox et al., 2020; Miech et al., 2018; Waltz et al., 2015).

However, there is surprisingly little research defining the concept of ‘change champion,’ and evaluating the impact of change champions on healthcare implementation efforts. Often research on champions is only descriptive in nature, lacking in detail, or the findings are embedded within broader, complex implementation efforts that cannot isolate the individual effect of the champions (Miech et al., 2018; Santos et al., 2022; Shea, 2021). To illustrate, two reviews on champions in healthcare implementation reported that the vast majority of articles only considered champions in terms of presence or absence (more than 90% of 199 articles (integrative review)(Miech et al., 2018), 71% of 35 articles (systematic review of quantitative research only)(Santos et al., 2022). Santos and colleagues’ (2022) systematic review of quantitative research related to healthcare champions reported that though champions were related to increased use of healthcare innovations at an organisational level (i.e. policies and processes), there was inconsistent evidence about whether champions were also related to improvements in provider’s attitudes and knowledge, use of innovations, and patient outcomes.

This lack of detailed research on champions is also observed in systematic reviews of organisational health literacy, all of which highlight the role of champions, but bear little detail about how to implement this strategy effectively. For example, there was no detail about who champions were, how champions were identified, what training they received, and what activities they engaged in as champions (Farmanova et al., 2018; Kaper et al., 2021; Lloyd et al., 2018). It is also possible that these reviews of organisational health literacy overlooked some articles relating to health literacy champions given the search terms they used. This oversight is important because the context of *health literacy* may be different to that of other healthcare champions. For example, health literacy initiatives can vary greatly in scale (e.g. within a specific department vs. initiatives that span across multiple services and sites), and often involve partnership across disciplines, professions, sectors, and community organisations (Sørensen et al., 2021). To capture the state of the literature relating to health literacy champions, this systematic review therefore aimed to identify the empirical and conceptual research that exists about health literacy champions, including descriptive accounts (e.g. of their roles, responsibilities, selection, and training), evaluations of training and implementation, and relevant models and theoretical frameworks. Although we can think about health literacy champions as including people who operate across sectors or services, and individuals who are exemplars of health literate practice, in this study we focus on health literacy champions who seek to improve the health literacy practices of other staff members within their organisation.

## Methods

### Protocol and registration

This systematic review was registered with the international Prospective Register of Systematic Reviews (PROSPERO) (CRD42022348816). No amendments to the registered protocol were required. The review is reported in accordance with the Preferred Reporting Items for Systematic reviews and Meta-analyses 2020 statement (Page et al., 2021).

### Review Question

What empirical and conceptual research exists about health literacy champions, including descriptive accounts, evaluations of champion effectiveness, evaluations of champion training and implementation, and relevant models and theoretical frameworks?

### Inclusion and exclusion criteria

For this review, we included English language articles published in peer-reviewed journals or published books that examined the concept of a health literacy champion. In line with the CFIR definition (CFIR Research Team-Center for Clinical Management Research, 2022), health literacy champions was taken to refer to staff within an organization who are involved in implementation, delivery, or provision of a health literacy initiative that seeks to improve the health literacy practices in other staff members. No limits were set for date of publication.

Studies were excluded if they met any of the following criteria:

1. Mentioned the concept of health literacy champion as a future direction only and text about champions was not directly related to aims, methods or results of the manuscript.
2. Concerned with mental health literacy champions only
3. Involved patient/community/peer-led education initiatives
4. Focused on health literacy improvement in patient or community populations, rather than improvement in health literacy practices in an organization

### Search Strategy

A database search of MEDLINE, Embase, CINAHL, Scopus, and PubMed was conducted on 8 August 2022. Article titles, abstract, and keywords were searched using the following search string, based on other reviews of healthcare champions (see for example, (Miech et al., 2018; Wood et al., 2020)):

(“health literacy” or “health literate”) AND (“champion*” or “change agent*” or “opinion leader*” or “liaison*” or “liason*” or “ambassador*” or “implementation leader*” or “emergent leader*” or “promoter*” or “advocate*”).

Where possible MeSH search terms were used.

Conference abstracts that appeared during the database searching were excluded but potentially relevant full text articles relating to these conference abstracts were identified and screened.

Systematic reviews on organizational health literacy were also identified and examined for any potentially relevant articles. Additionally, a snowballing approach was used which involved searching the reference lists and citations (‘cited by’ in Google Scholar) of eligible articles.

### Study selection process

After duplicates were removed, titles and abstracts were independently screened by two authors (JA and MZ) for full text screening. All full texts were also independently screened for inclusion by these authors (JA and MZ). Any disagreements during this process were resolved through discussion between study authors.

### Data extraction and synthesis

Data for each article that met the inclusion criteria were independently extracted by two authors (JA & MZ). Extracted data was compared by the two authors and any differences were resolved through discussion with author KM. Data extracted included year of publication, aims, study setting and design, interventions implemented, details about the health literacy champions (role, responsibilities, selection, training and effectiveness, and any potential facilitators or barriers to successful championship. Following data extraction, patterns across the data were explored and synthesised in narrative form (Popay et al., 2006). Given the lack of quantitative data and limited detail, even in qualitative research, we did not seek to undertake subgroup analyses or sensitivity analyses. Findings about effectiveness were only synthesised for studies of low to moderate risk of bias.

### Quality appraisal

All full texts included in the data extraction process were assessed for risk of bias by two authors (JA and MZ) using standardized critical appraisal tools from JBI (https://jbi.global/critical-appraisal-tools). Depending on the study design, different JBI critical appraisal tools were used. These included the Checklist for Qualitative Research, and the Checklist for Quasi-Experimental Studies. All Text and Opinion articles were considered high risk of bias. For case study designs, as there was no JBI critical appraisal tool for these study types, these studies were assessed as high risk of bias. When articles contained multiple study design components, a checklist was utilized for all study designs relevant to health literacy champions. Using these tools, studies were categorised as: low risk of bias if most criteria were fulfilled and done well, moderate risk of bias if some of the criteria were fulfilled, or high risk of bias if most criteria were not done or done poorly. Discrepancies in ratings between the two authors were resolved through discussion.

## Results

### Study Details

We retrieved 1149 articles from the database searches, and 18 from additional search methods (Figure 1). After removal of duplicates and screening by title and abstract, 55 full-text articles were screened for full-text inclusion. Articles were excluded if champions were community members rather than staff (n = 7), if they reported on health literacy improvement in patient or community populations, rather than improvement in health literacy practices in an organization (n = 11), or if the review’s definition of champion was otherwise not met i.e. the champion did not influence others within their organisation (n = 20). Seventeen articles met our inclusion criteria and were included in the final synthesis.

**Figure 1.**
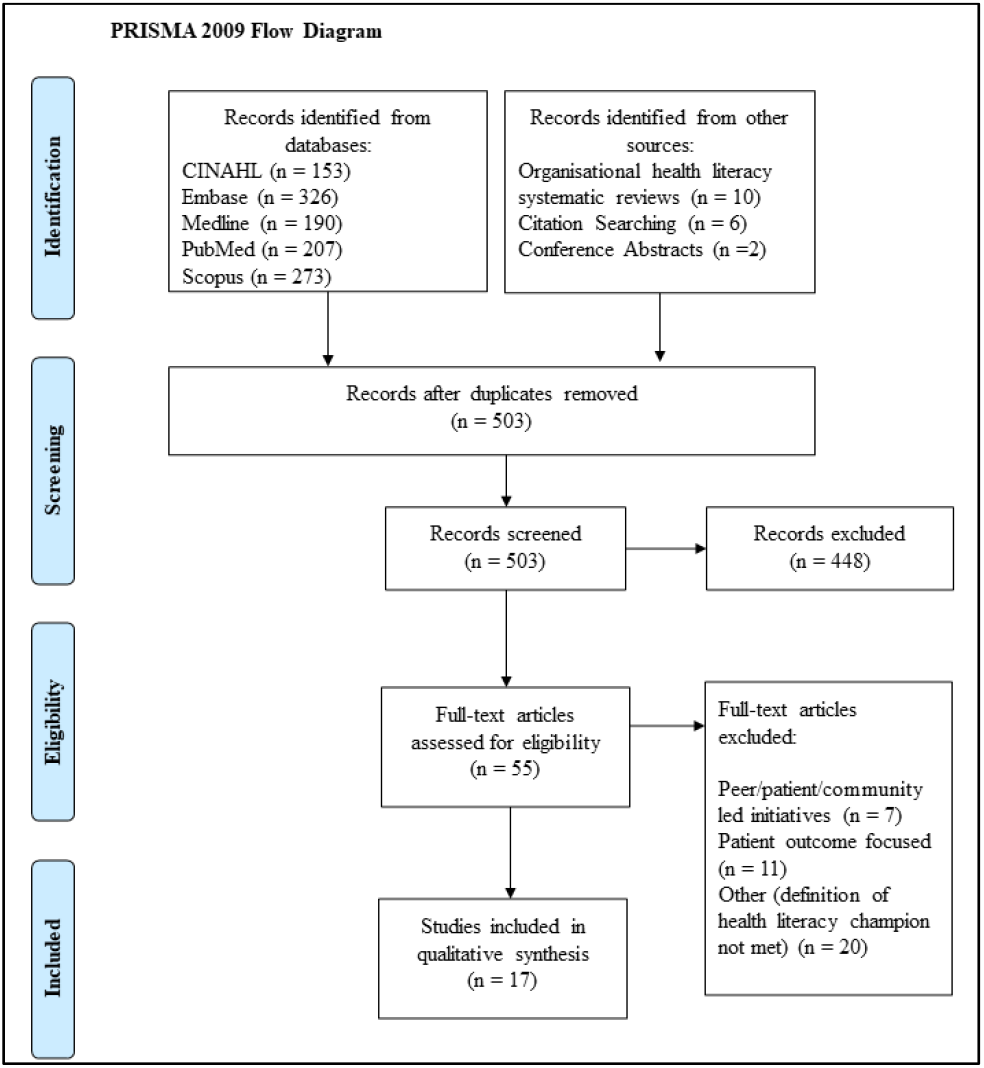
Preferred reporting items for systematic reviews and meta-analyses (PRISMA) flow diagram of the study selection process. Adapted from Moher et al., (2009)

### Study characteristics and risk of bias assessment

The 17 studies identified in the review are described in Table 1. With two exceptions, health literacy champions were not the primary focus of the included research articles and were mentioned as one aspect of implementation of a health literacy intervention or initiative. Only two articles focused primarily on health literacy champions (Brach et al., 2014; Sørensen, 2021). These were both of the ‘Text and Opinion’ article type, with one providing additional case studies (Sørensen, 2021). Overall, nine articles adopted a case study design and provided an account of how organisational health literacy was introduced in an organisation, with health literacy champions playing some part in this process (all high risk of bias). Four articles reported qualitative research investigating how health literacy practices (Adsul et al., 2017; Carol J. Howe et al., 2020) or tools (Kaper et al., 2019; Mabachi et al., 2016) had been implemented within an organisation (three low and one moderate risk of bias). Two studies used quasi-experimental designs to evaluate the implementation of a health literacy intervention in clinical settings (Morrison et al., 2021; O’Neal et al., 2013), although effects of champions were not isolated from the broader intervention (one low, one high risk of bias). Five articles were categorised as ‘Text and Opinion’ (two without accompanying case studies) and primarily provided a conceptual account of how organisations can improve their health literacy practices.

**Table 1.** Characteristics of included studies.

| First author (year) | Setting | Study aims | Intervention | Who were the HL champions? | HL champion role, responsibilities, activities performed | Risk of Bias |
| --- | --- | --- | --- | --- | --- | --- |
| **Qualitative studies** |  |  |  |  |  |  |
| Adsul (2017) | US low-resourced health care organisations | Identify how health care organizations serving underserved populations adopt and implement health literacy related changes | Health literacy quality improvement program | Staff who were familiar with all aspects of the organization, recognized need for improvements in patient care. | Lead implementation/ facilitate change of health-literacy practices in organization ('program champion') | Moderate |
| Howe (2020) | US (Texas) hospitals | Describe key organisational leaders' and clinicians' perspectives on how health care systems were adopting health literate policies and practices that address the 10 attributes of a health literate healthcare organization | None described | Out of 13 services, only 1 had an appointed dedicated health literacy position; 1 additional example of a clinical nurse (frontline worker) who was an emergent champion | • Involve patients in information processes e.g. development and review of booklets<br>• Delivering patient education in a creative manner e.g. using smartphones and tablets | Low |
| Kaper (2019) | Ireland, Netherlands | Assess the implementation fidelity, moderators (barriers and facilitators), and long-term impact of organisational HL interventions in hospitals | Organisational HL intervention, including HL audit, implementation coordinators, project committees | Appointed staff including senior nurses, policy advisor, communications officer, communication consultants | • Coordinate implementation process<br>• Embed health literacy in working procedures and professional practice | Low |
| Mabachi (2019) | US primary care practices | 1. Examine utility of the health literacy universal precautions toolkit for primary care practices<br>2. Identify possible refinements that would enhance the value of the Toolkit as a resource for primary care practices. | Universal precautions toolkit with support from contracted external health literacy experts | 1. Appointed health literacy team leaders<br>2. Practice leaders (e.g. management level) | • Team leaders: oversee implementation<br>• Practice leaders: directly involved in planning and implementation, publicly empower the health literacy team to carry out project activities | Low |
| **Quasi experimental** |  |  |  |  |  |  |
| Morrison (2021) | US emergency departments | Improve the distribution, satisfaction, and understanding of asthma education and discharge instructions in parents of children with asthma exacerbation in the ED | Process maps to understand workflows and inform the health literacy initiative (4 stages: education, integration, refresher and sustainment) | Physician and education expert, 4 ED nurse champions and a quality improvement specialist. | • Develop an implementation strategy and new materials<br>• Trial new materials/strategy<br>• Work closely with IT and quality improvement staff<br>• peer to peer education (nurses) about educational materials and health literacy-focused communication | Low |
| O'Neal (2013) | US community pharmacies | 1. Pilot, evaluate and adapt Pharmacy Health Literacy Assessment Tool<br>2. Describe use of health literacy practices from patient, staff, and independent auditor perspectives<br>3. Evaluate the effect of a low-intensity educational health literacy awareness program<br>4. Identify opportunities to improve health literacy sensitive practices in the community pharmacy environment. | AHRQ pharmacy HL assessment tool; 2 hour HL literacy training workshop for pharmacists delivered by nationally recognised expert | Pharmacy staff who attended the training | Initiate literacy sensitive practices at their pharmacy (the extent of this was up to the champion) | High |
| Case studies |  |  |  |  |  |  |
| Allott (2018) | Health and community services in Australia | Describe how a 'system approach' to health literacy responsiveness created change in regional health and community services | 1. Health literacy training course (8 months)<br>2. Executive level workshop<br>3. Community of practice (meetings and networks)<br>4. Alliance to facilitate peer support, problem solving, and regional action | Not specific if emergent or appointed. Some champions had attended the training course | Champion health literacy principles and practices within the organisation | High |
| Brach (2017) | US health care organizations that have pioneered a systematic approach to health literacy | Explore the process for organizations to become health literate health care organizations | None described | HL champions can be:<br>1. Organizational leaders (e.g. CEO's)<br>2. Formal health literacy liaisons (appointed)<br>3. Staff who have informally become standard bearers for health literacy in the course of their jobs (emergent) | Organizational leader:<br>• Initiate research<br>• Engage other organizational leaders<br>• Establish a HL taskforce<br>• Implement policies in hospitals<br>Formal HL liaison:<br>• Ensure health information is easy to access, understand, and use<br>• Ensure system-wide processes take health literacy into account<br>• Promote system-wide health literacy standards<br>• Develop a system-wide strategic plan for health literacy<br>Emergent:<br>• Advocate through passion for improving patient education and written materials<br>• Raise health literacy to the top levels of an organization | High |
| Briglia (2015) | US federally qualified health centre | Provide steps for becoming a health literate organisation | None described | Contracted health literacy program manager | • Lead HL initiative<br>• Create and communicate a change vision, generate staff awareness about HL<br>• Establish a HL task force<br>• Implement staff training and agency-wide HL policies<br>• Ensure all patient materials meet HL guidelines | High |
| Erikson (2019) | US health services (state level) | Outline how an adult basic education coalition developed a state-wide health literacy coalition | None described | Emergent champion was an influential physician who joined the board of directors for Wisconsin Literacy Inc | • Deliver presentations on HL at state and national conferences<br>• Create opportunities and influence conversations in health care settings | High |
| Finlay (2019) | Acute and community health services in a regional area of Australia | Report on outcomes of a project (Gippsland Health Literacy Project) designed to educate local health services staff about health literacy and provide tools and techniques for health literacy implementation in service delivery | 1. An introductory health literacy forum<br>2. A 2-day health literacy short course<br>3. Training in the use of specific health literacy tools<br>4. Completion by participants of individual 'plan do study act' | Not described | Not described | High |
| O'Neill (2019) | US health services | Provide a case study/example model of health literacy change in a health system | None described | Emergent:<br>1. Nurse-educator from Nursing and Professional Development & Education department<br>2. Chief learning Officer<br>Identified:<br>3. Medical Director of Patient & Family Health Education | Nurse:<br>• Campaign for the issue of health literacy in leadership venues e.g. nursing councils and service line leader meetings<br>• Organise Health Literacy Awareness Month events<br>• Campaign to department director and Chief Learning Officer (member of CEO roundtable)<br>• Assess HL practices, present findings to senior leadership<br>Chief learning officer:<br>• Develop system-wide organisational processes to create uniform messaging about health literacy<br>• Consolidate education content vendors across the organisation<br>• Allocate more personnel resource to support these changes and future efforts<br>• Establish MD as additional co-advocate to increase credibility, perspective and relatability to conversations about health literacy practices<br>Medical director:<br>• Form a governance group including consumers | High |
| Shoemaker (2013) | US pharmacies | Identify facilitators and barriers to uptake and implementation of AHRQ's health literacy tools | AHRQ pharmacy health literacy tools | 1. Part-time pharmacy staff (e.g., clinical faculty, students)<br>2. Pharmacy staff with designated responsibilities besides dispensing (e.g., patient care)<br>3. Full-time Director of Pharmacy Operations | Implement the tool | High |
| Sørensen (2021) | Health organisations (international focus) | Explore health literacy championship and how health literacy champions are characterized and nurtured as change-agents for the development of health literate organizations, settings and societies. | Gives example of Health Literacy Champion Process (for Nebraska LHD), in which an organisation can apply to be a designated Health Literacy Champion to incentivise implementation of HL practices | Describes champions as leaders and/or decision-makers in the organization who have the influence needed to approve or put the plan into action.<br>Distinguishes this role from allies (supporters), and workgroup members (day to day HL activities) (CDC case study). Champions may be internal or external to the organisation.<br>Characteristics include patience, endurance, and long-term vision ("enthusiastically and relentlessly defends and fights for the cause of health literacy to the benefit of people and societies at large") | • 'Induce' and develop the change in organisational thinking<br>• Explain the necessity to perform a change of practice<br>• Generate general or public interest to effect change | High |
| Vellar (2016) | Australian regional health service | Describe how health service embedded health literacy principles over a 3-year period | Three phases to implementation of a HL Framework:<br>1. Literature review and critical reflection on critical incidences<br>2. Organisational commitment and consultation process<br>3. Piloting health literacy strategies for system-wide improvements, including HL ambassador (HLA) program that trains staff to lead their teams on how to partner with consumers to develop plain English resources | 1. Appointed health literacy ambassador<br>2. Senior executive leadership | HL ambassador: lead development of plain English resources for patients<br>Senior executive leadership: advocate for HL within their level of influence and advocate for HL as a quality and safety issue | High |
| **Text and opinion** |  |  |  |  |  |  |
| Brach (2013) | US health services | Describe physician's role in a creating a health literate organisation | None described | Physicians within a health organisation | • Promote integration of health literacy into strategic and operational planning, evaluation and quality improvement<br>• Insist on HL training for all staff, and system redesign to meet needs of patients at all HL levels<br>• Install mechanisms to obtain input from patients to improve services<br>• Direct resources to where misunderstandings would have the most severe consequences<br>• Promote price transparency to make out of pocket costs easy to understand and timely | High |
| Erlen (2004) | US | Provide an overview of functional health illiteracy, identify related ethical concerns, and discuss selected, relevant nursing implications | None described | Nurses | • Increase awareness of health literacy amongst other health professionals<br>• Develop protocol to assess individual health literacy<br>• Direct healthcare staff to health literacy research articles and HL measures<br>• Advocate for health literate patient education materials<br>• Facilitate decision making: explain difficult information and translate technical language into language that patients understand (though this is pitched as working directly with patients) | High |

### Outcomes

#### Who were the health literacy champions?

Health literacy champions were most often described in terms of their professional role. For example, champions included nurses (Erlen, 2004; Kaper et al., 2019; Morrison et al., 2021; O’Neill, 2019), physicians (Brach et al., 2014; Erikson et al., 2019; Morrison et al., 2021), pharmacists (O’Neal et al., 2013; Shoemaker et al., 2013), medical residents (Shoemaker et al., 2013), and staff involved in policy, communication and quality improvement (Kaper et al., 2019; Morrison et al., 2021). Another important group were champions in positions of leadership, including at the executive level (Brach, 2017; Mabachi et al., 2016; Shoemaker et al., 2013; Sørensen, 2021). Two studies described champions who were consultants or externally contracted staff with expertise in health literacy (Briglia et al., 2015; Kaper et al., 2019). Champions were also variously described ‘emergent’ (Erikson et al., 2019; Sørensen, 2021) (i.e. staff who take on a champion role of their own accord due to their high commitment to the cause), or as staff ‘appointed’ to a champion role (Briglia et al., 2015; Kaper et al., 2019; Mabachi et al., 2016). Sometimes emergent champions worked in services that did not initially value or engage in health literacy practices e.g. (Erikson et al., 2019; O’Neill, 2019).

However, this was not always the case. For example, Brach (2017) discussed that CEO-level staff often became health literacy champions in part due to alignment with the organization’s mission and goals.

If more than one champion was present, a combination of both emergent and appointed champions was often present (Brach, 2017; C. J. Howe et al., 2020; O’Neill, 2019; Shoemaker et al., 2013; Vellar et al., 2017). Typically champions in a senior leadership position were ‘emergent’, whereas staff on the ground were either emergent or appointed (Brach, 2017; Mabachi et al., 2016; O’Neill, 2019; Shoemaker et al., 2013; Vellar et al., 2017).

#### Champion training

Five studies described health literacy training programs for champions, which ranged in duration from a single 2-hour workshop (O’Neal et al., 2013; Vellar et al., 2017) through to eight months of ongoing training (Allott et al., 2018). Two studies described a continuation of learning through ongoing mentoring and collaborative support from other champions (Allott et al., 2018; Vellar et al., 2017). Of the five studies that included training, only two mentioned specific training in implementation skills in addition to general health literacy knowledge and skills (Finlay et al., 2019; Morrison et al., 2021). Kaper and colleagues (2019) described how implementation skills were supplemented by ‘implementation coordinators’.

More than half of the articles (n=9) did not describe any form of training for health literacy champions, with most of these focussing on emergent champions.

#### Health literacy champion activities, roles and responsibilities

Key activities (or roles and responsibilities) fell broadly into three categories: increasing health literacy awareness and organisational commitment; and changing strategic and operational planning; and influencing frontline health literacy practices.

Generating awareness about health literacy was typically focused within the organisation but occasionally extended beyond (Briglia et al., 2015; Erikson et al., 2019; Erlen, 2004; O’Neill, 2019; Sørensen, 2021). This encompassed communicating the change vision and advocating health literacy to organisational leaders. Three articles described that health literacy champions could seek to influence other organisational leaders to support health literacy initiatives or become health literacy champions themselves (Brach, 2017; Erikson et al., 2019; O’Neill, 2019).

A second key activity was influencing strategic and operational planning (Brach, 2017; Brach et al., 2014; Briglia et al., 2015; Kaper et al., 2019; Mabachi et al., 2016; O’Neill, 2019). On a broad level this was described as changes to organisational policy (Brach, 2017; Briglia et al., 2015) and processes (Brach, 2017; Kaper et al., 2019). For example, for three studies this explicitly involved linking health literacy to existing quality improvement processes and IT services (Brach et al., 2014; Morrison et al., 2021; Vellar et al., 2017).

Several articles highlighted the role of champions in influencing frontline health literacy practices. Six articles described the champion as ensuring that the organisation’s health information materials met health literacy guidelines (Brach, 2017; Briglia et al., 2015; Erlen, 2004; C. J. Howe et al., 2020; Morrison et al., 2021; Vellar et al., 2017). Three studies described how champions implemented health literacy training for staff (Brach et al., 2014; Briglia et al., 2015; Morrison et al., 2021); three described establishing a health literacy task force, working group, or committee (Brach, 2017; Briglia et al., 2015; O’Neill, 2019) (though with little detail about the aims of these groups); and three described advocacy and implementation of mechanisms to increase consumer engagement in the organisation’s practices (Brach et al., 2014; C. J. Howe et al., 2020; O’Neill, 2019). Other activities included assessment of organisational health literacy practices (O’Neal et al., 2013; O’Neill, 2019; Shoemaker et al., 2013), or of individual (patient) health literacy (Erlen, 2004).

Two studies did not provide specific details and simply alluded to the champions leading implementation and advocating for health literacy (Adsul et al., 2017; Allott et al., 2018).

#### Potential facilitators and barriers to successful championship

Several studies identified the importance of support and commitment to health literacy initiatives from executive leadership (Allott et al., 2018; Brach, 2017; Finlay et al., 2019; C. J. Howe et al., 2020; Mabachi et al., 2016; Sørensen, 2021). Many also emphasised that health literacy champions cannot act in isolation, and recommended multiple champions at varying levels within the organisation (Brach, 2017; C. J. Howe et al., 2020; Sørensen, 2021; Vellar et al., 2017). Further, champions can be supported by other groups within the organisation; Sørensen (2021) describes the CDC cast study which depicts champions as working in unison with allies (who provide support/vision), and workgroup members (day to day planning and coordination).

Some studies described the importance of organisational awareness and commitment to health literacy *before* appointing health literacy champions (Mabachi et al., 2016), for supportive policies and infrastructure to be in place (Brach et al., 2014), and for a culture that fosters innovation and quality improvement (Sørensen, 2021).

Lastly, limited resources, dedicated personnel and funding were often identified as barriers to effective health literacy champions (Brach, 2017; C. J. Howe et al., 2020; Shoemaker et al., 2013).

For appointed champions, bolstering commitment to health literacy may also be important. The authors of two studies proposed several examples of strategies that could strengthen this commitment: personal invitation to champion health literacy from a trusted source e.g. academic institution; awards and other incentives; and aligning champion activities with other goals (such as meeting residency requirements) (Shoemaker et al., 2013; Sørensen, 2021). Shoemaker and colleagues (2013) also suggested that providing ongoing support from health literacy experts helped strengthen the commitment of health literacy champions.

#### Effectiveness

Overall, five articles with low to moderate risk of bias reported on the effectiveness of champions. This included four qualitative studies and one quasi-experimental study. Three reported positive effects (Adsul et al., 2017; C. J. Howe et al., 2020; Morrison et al., 2021). The remaining two articles reported neutral effects of health literacy champions (e.g. the champion was only one component of the health literacy initiative and was not identified as a critical factor) (Kaper et al., 2019; Mabachi et al., 2016). Two of the three studies reporting positive effects involved emergent champions, and the third study did not report this characteristic; by comparison, the two studies reporting neutral effects involved appointed champions.

The quasi-experimental study explored a health literacy initiative to improve asthma education in a US emergency department (Morrison et al., 2021). Champions were only one component of this initiative, and their unique effects were not reported. Study authors reported an increase in families receiving asthma education over a 12 month period for written (28% to 52%) and video materials (0% to 32%), although no statistical analysis was performed. The intervention did not result in changes to emergency department length of stay, length of discharge, or 30 day revisit rates.

## Discussion

We identified 17 articles related to health literacy champions that were generally of high risk of bias. These articles provided only very limited detail about champions, in part because the articles focused on multi-component implementation efforts. Champions included staff on the ground (e.g. nurses, physicians, pharmacists), in administrative or management roles (e.g. quality improvement, senior nurses, communication), and in executive leadership roles. Few studies described training for health literacy champions, and those that did provided little detail. Key champion activities related to increasing organisational awareness and commitment to health literacy, influencing strategic and operational planning, and influencing frontline health literacy practices. The most frequently described influence on frontline practices was to ensure that the organisation’s health information met health literacy guidelines. Articles recommended having multiple champions at varying levels within the organisation, including the executive level. Limited funding and resources were identified as key barriers for health literacy champions. Three of five studies with low to moderate risk of bias reported that champions may enhance implementation of health literacy initiatives, though further work is needed to isolate the effect of champions from other implementation strategies.

These findings highlight a clear lack of a foundational, rigorous evidence base that health services can draw upon to inform their health literacy champion roles, programs, and training. Champion research in the broader healthcare literature faces similar issues. For example, most studies only report on the presence or absence of a champion, and do not separate the unique effects of champions from broader multi-component implementation efforts (Lennox et al., 2020; Miech et al., 2018; Shea, 2021). To build a stronger evidence base, health literacy champion research must include experimental study designs and process evaluations that focus specifically on the champions themselves. This must also be accompanied by more detailed reporting (e.g. staff involved, training and expected roles). Over time we may then develop a better understanding of why a given health literacy champion initiative may or may not have worked (Powell et al., 2019).

This review did identify some promising directions for health services looking to establish health literacy champions. Notably, several articles described having *multiple champions* working simultaneously in a coordinated way, with some champions being at the executive or senior leadership levels. This finding is consistent with other systematic reviews of health care champions, which reported that these ‘network’ structures may be more effective than solo champions (Miech et al., 2018). Interestingly, this review identified a mix of ‘top-down’ health literacy champion networks, such as the CDC model of ‘champions,’ ‘allies,’ and ‘workgroup members’ described by Sørensen (2021); and less hierarchical approaches such as Allott and colleagues’ (2018) champions who were nested within a community of practice and alliance network that encouraged collaboration and problem-solving with other champions. The Health Literacy Hub in Western Sydney is another useful example of how a community of practice model can support champions. Over a five year period, the Hub has grown to more than 1300 members, providing them with health literacy information and tools, and connecting with members via seminars, mailing lists, targeted training, and partnerships or consultation projects (Muscat et al., 2023). The initiative emphasises the role of trust, co-creation, and partnership synergy in creating an effective and sustainable community of practice. Further work is needed to inform how health services can create their own sustainable networks of health literacy champions that build staff health literacy knowledge and skills, across a variety of health service settings and organisational structures.

Current organisational health literacy resources lack detailed guidance about how to identify, prepare, and support champions. For example, champions are not mentioned in the organisational health literacy responsiveness framework (Trezona et al., 2017) and the ‘Ten attributes of a health-literate organisation’ only briefly mentions the need to ‘cultivate health literacy champions throughout an organisation’ (Brach et al., 2012). The CDC provides some greater detail, advocating that a first step to improving organisational health literacy practices is to establish champions, allies and workgroup members (Centers for Disease Control and Prevention, 2022). Although a stronger evidence base is needed for concrete recommendations, these resources could guide health services to reflect on who their champions might be, the scope of their roles, expected output, and the kind of incentives, training, or support they need. Given this review highlighted that the commitment of champions may waver, the resource could also include reflection on each champion’s personal motivation for improving health literacy, and potential incentives to maintain their commitment.

The strengths of this study were that a wide range of ‘champion’ search terms were included, across multiple databases. Limitations are that only English-language articles were captured. It is also worth noting that there is also some overlap between the concepts of ‘leaders’ and ‘champions’ (Damschroder et al., 2022). Studies that reported solely on leadership support for health literacy are not captured in this review. Limitations of the primary studies were that they generally had high risk of bias and champions were often not described in detail. As a result, this review cannot provide definitive conclusions about whether champions were effective, nor in which contexts.

Despite the potential positive impacts of health literacy champions, this review suggests that more high-quality research on health literacy champions is needed. As a first step, quality can be improved through more comprehensive reporting on health literacy champions, including who the champions are, the training they received, and the tasks they carried out. Experimental and process evaluation research across multiple sites will also contribute valuable insights into this implementation strategy. Engaging multiple champions at varying levels within the organisation, including the executive level, is a promising future direction for this area of research.

## Data Availability

Data available within the article or its supplementary materials

## Data availability

Data available within the article or its supplementary materials

## Acknowledgements

Not applicable.

## Notes

**Funding** JA is supported by the National Health and Medical Research Council (Grant number: 2017278).

### Competing Interest Statement

The authors have declared no competing interest.

### Funding Statement

JA is supported by the National Health and Medical Research Council (Grant number: 2017278).

